# Collaborative Cohort of Cohorts for COVID-19 Research (C4R) Study: Study Design

**DOI:** 10.1101/2021.03.19.21253986

**Authors:** Elizabeth C Oelsner, Norrina Bai Allen, Tauqeer Ali, Pramod Anugu, Howard Andrews, Alyssa Asaro, Pallavi P Balte, R Graham Barr, Alain G Bertoni, Jessica Bon, Rebekah Boyle, Arunee A Chang, Grace Chen, Shelley A Cole, Josef Coresh, Elaine Cornell, Adolfo Correa, David Couper, Mary Cushman, Ryan T Demmer, Mitchell S. V. Elkind, Aaron R Folsom, Amanda M Fretts, Kelley Pettee Gabriel, Linda Gallo, Jose Gutierrez, MeiLan K. Han, Joel M Henderson, Virginia J Howard, Carmen R Isasi, David R Jacobs, Suzanne E Judd, Debora Kamin Mukaz, Alka M Kanaya, Namratha R Kandula, Robert Kaplan, Akshaya Krishnaswamy, Gregory L Kinney, Anna Kucharska-Newton, Joyce S. Lee, Cora E Lewis, Deborah A. Levine, Emily B. Levitan, Bruce Levy, Barry Make, Kimberly Malloy, Jennifer J Manly, Katie A Meyer, Yuan-I Min, Matthew Moll, Wendy C. Moore, Dave Mauger, Victor E. Ortega, Priya Palta, Monica M Parker, Wanda Phipatanakul, Wendy Post, Bruce M Psaty, Elizabeth A Regan, Kimberly Ring, Véronique L. Roger, Jerome I Rotter, Tatjana Rundek, Ralph L. Sacco, Michael Schembri, David A. Schwartz, Sudha Seshadri, James M Shikany, Mario Sims, Karen D Hinckley Stukovsky, Gregory A Talavera, Russell P Tracy, Jason G Umans, Ramachandran S Vasan, Karol Watson, Sally E. Wenzel, Karen Winters, Prescott G. Woodruff, Vanessa Xanthakis, Ying Zhang, Yiyi Zhang, For the C4R Investigators

## Abstract

The Collaborative Cohort of Cohorts for COVID-19 Research (C4R) is a national prospective study of adults at risk for coronavirus disease 2019 (COVID-19) comprising 14 established United States (US) prospective cohort studies. For decades, C4R cohorts have collected extensive data on clinical and subclinical diseases and their risk factors, including behavior, cognition, biomarkers, and social determinants of health. C4R will link this pre-COVID phenotyping to information on SARS-CoV-2 infection and acute and post-acute COVID-related illness. C4R is largely population-based, has an age range of 18-108 years, and broadly reflects the racial, ethnic, socioeconomic, and geographic diversity of the US. C4R is ascertaining severe acute respiratory syndrome coronavirus 2 (SARS-CoV-2) infection and COVID-19 illness using standardized questionnaires, ascertainment of COVID-related hospitalizations and deaths, and a SARS-CoV-2 serosurvey via dried blood spots. Master protocols leverage existing robust retention rates for telephone and in-person examinations, and high-quality events surveillance. Extensive pre-pandemic data minimize referral, survival, and recall bias. Data are being harmonized with research-quality phenotyping unmatched by clinical and survey-based studies; these will be pooled and shared widely to expedite collaboration and scientific findings. This unique resource will allow evaluation of risk and resilience factors for COVID-19 severity and outcomes, including post-acute sequelae, and assessment of the social and behavioral impact of the pandemic on long-term trajectories of health and aging.

---

The adverse effects of the coronavirus disease 2019 (COVID-19) pandemic on United States (US) health, economy, and society are widespread and will likely continue well beyond the initial waves of infections (1). Lack of preparedness and inadequate implementation and uptake of standard public health interventions in the US has already contributed to over 23 million cases, one million hospitalizations and over 500,000 deaths from COVID-19 (2, 2), making COVID-19 the third-leading cause of death in the United States in 2020 and the second-leading cause of death in those over 85 years of age (4, 2). Furthermore, prolonged symptoms and clinical abnormalities are observed in some COVID-19 survivors, raising concerns that post-acute sequelae of COVID-19 could pose an additional long-term health burden (6).

Epidemiologists have marshalled the strengths of numerous complementary study designs to identify the incidence and major clinical and socio-demographic risk factors for COVID-19 illness, as well as to describe post-COVID-19 outcomes. In particular, case-based registries and large-scale electronic health record (EHR)- and health systems-based cohorts provided critical early insights into disease susceptibility and short- and long-term sequelae. Among these were findings that socio-economic disadvantage (5, 7-9) and pre-existing clinical conditions, such as obesity, heart conditions, or lung disease (10-19), are associated with greater risk of severe illness.

Nonetheless, clinical and survey databases pose several problems for COVID-19 epidemiology. Clinical case series lack rigorous control groups, have non-standardized, limited data collection, and are subject to ascertainment biases – including, but not limited to, reduced health care access and quality among vulnerable communities. EHRs typically lack detailed information on health-related behaviors, such as smoking, so that controlling for confounders is challenging. Moreover, in the course of usual clinical care, clinically actionable diagnostic testing is performed for sick persons, but not well persons; hence, subclinical disease is not well detected, and genomic and other mechanistic biomarkers are generally lacking. Although inception cohorts with longitudinal follow-up of clinically ascertained cases of COVID-19 cases can address some of these knowledge gaps, survival bias, recall biases, and non-randomly missing data regarding pre-COVID health and behaviors are inevitable. In this context, strong assumptions are required to define phenotypes identified in COVID-19 survivors (e.g., fibrotic lung disease) as “sequelae” when they may have been present prior to the pandemic, and actually be antecedent risk factors or effect modifiers.

The Collaborative Cohort of Cohorts for COVID-19 Research (C4R) was established as a national, prospective study of adults at risk for incident COVID-19 that is relatively free of referral, survival, and recall biases. C4R includes fourteen US prospective cohort studies that, collectively, constitute a large, well-characterized, population-based sample that ranges in age from young adults to centenarians, and reflects the racial, ethnic, socioeconomic, and geographic diversity of the US. Using standardized protocols, C4R is aggressively attempting full ascertainment of SARS-CoV-2 infection and COVID-19 illness across all cohorts. C4R offers the additional major advantages of standardized data collection protocols, including high-quality clinical events surveillance dating back as far as 1971 in some studies, and robust retention rates.

For decades, the C4R cohorts have collected extensive longitudinal data on clinical and subclinical disease, behaviors, cognition, biomarkers, and social determinants of health. C4R will link this “pre-COVID” phenotyping to information on SARS-CoV-2 infection and acute and post-acute COVID-related illness. The integration of antecedent and illness-related data will provide a unique opportunity to understand mechanisms and modifiers of risk and resilience for SARS-CoV-2 infection and adverse COVID-19 outcomes. C4R will also support comparisons of longitudinal changes in health measures over the course of the pandemic in persons with varying degrees of COVID-19 severity. Furthermore, the availability of well-characterized participants unaffected by COVID-19 will allow the assessment and differentiation of the effects of infection, illness, and pandemic-related social, economic, and behavioral changes.

Overall, C4R aims to provide a valuable scientific resource to (1) evaluate risk and resilience factors for adverse COVID-19 outcomes, including severe COVID-19 illness and long-term complications, (2) assess the social and behavioral impact of the COVID-19 pandemic on long-term outcomes and trajectories of health and disease, and (3) examine disparities in COVID-19 risk and outcomes according to race, ethnicity, geography, and other social determinants of health.

## METHODS

### Cohort of cohorts

Fourteen prospective cohorts are collaborating in C4R (**Table 1**). Eight of the cohorts were designed to study cardiovascular disease epidemiology: Atherosclerosis Risk in Communities (ARIC) Study (20), Coronary Artery Risk Development in Young Adults (CARDIA) Study (21), Framingham Heart Study (FHS) (22), Hispanic Community Health Study/Study of Latinos (HCHS/SOL) (23-25), Jackson Heart Study (JHS) (26-28), Mediators of Atherosclerosis in South Asians Living in America (MASALA) Study (29, 2), Multi-Ethnic Study of Atherosclerosis (MESA) (31), and the Strong Heart Study (SHS) (32, 2). These cohorts generally recruited population-based samples, although only three (ARIC, CARDIA, FHS, HCHS/SOL) used representational sampling techniques at some or all sites. Four of the cardiovascular studies (ARIC, CARDIA, FHS, MESA) recruited multi-racial participants, and four were designed to study primarily specific race or ethnic groups (Hispanic/Latino participants in HCHS/SOL, Black participants in JHS, South Asian participants in MASALA, American Indian participants in SHS). Four multi-ethnic cohorts were established to study respiratory epidemiology: the Genetic Epidemiology of COPD (COPDGene) Study (34) and the SubPopulations and InteRmediate Outcome Measures in COPD Study (SPIROMICS) (35) were established as longitudinal case-control studies of cigarette smokers with and without COPD; Prevent Pulmonary Fibrosis (PrePF) is a study of early and established interstitial lung disease; and, the Severe Asthma Research Program (SARP) is a study of the entire range of mild to severe asthma, enriched for severe disease (36). Two studies – the Northern Manhattan Study (NOMAS) and the REasons for Geographic and Racial Differences in Stroke (REGARDS) – were established to study primarily neurological outcomes, including stroke and cognition. NOMAS is a multi-ethnic community study (37) and REGARDS is a biracial (non-Hispanic Black, White) national sample of the continental US that oversampled Black people and those residing in the southeast (38).

**Table 1.**
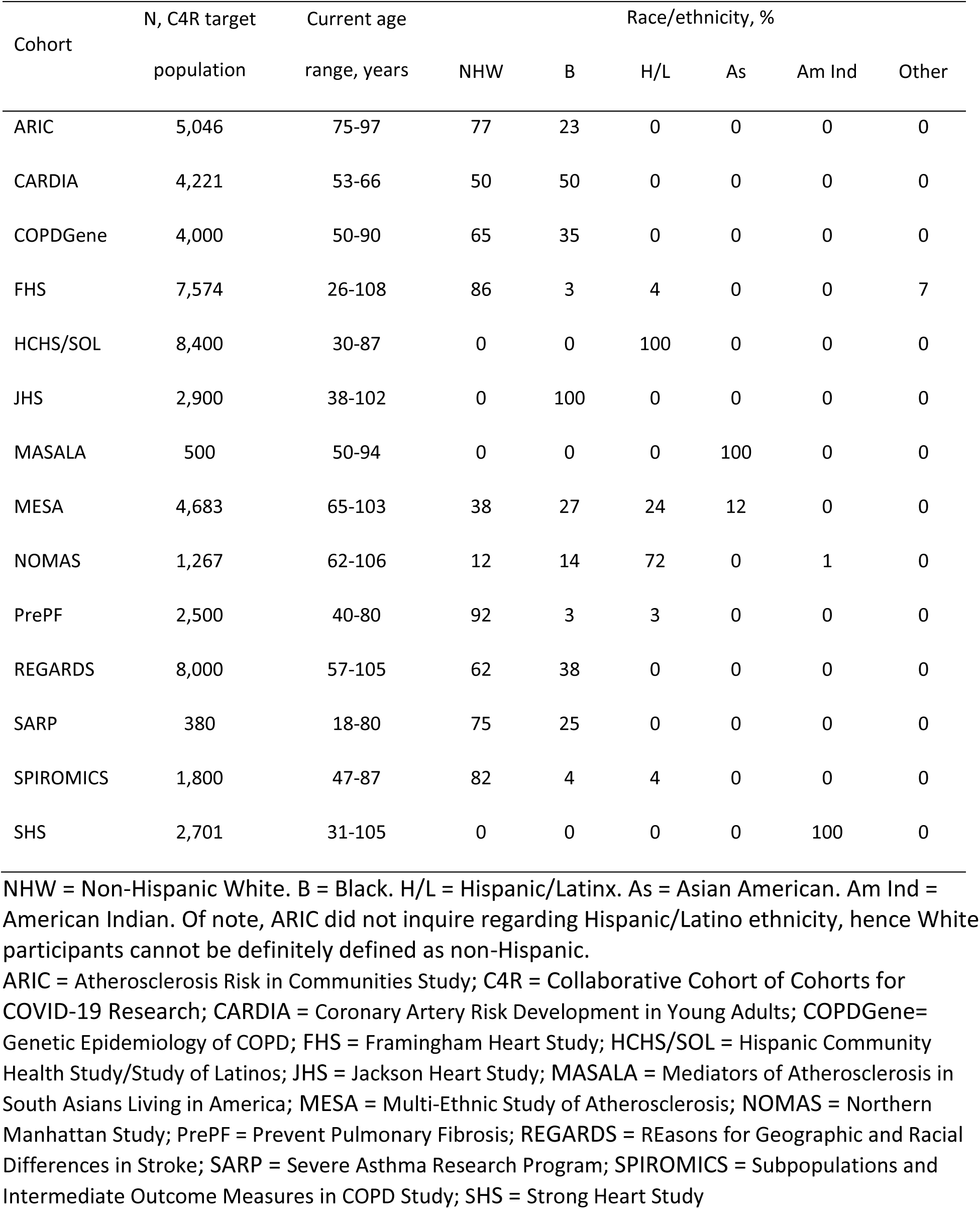
Characteristics of participants in C4R cohorts on March 1, 2020.

| Cohort | N, C4R target | Current age | Race/ethnicity, % |  |  |  |  |  |
| --- | --- | --- | --- | --- | --- | --- | --- | --- |
|  | population | range, years | NHW | B | H/L | As | Am Ind | Other |
| ARIC | 5,046 | 75-97 | 77 | 23 | 0 | 0 | 0 | 0 |
| CARDIA | 4,221 | 53-66 | 50 | 50 | 0 | 0 | 0 | 0 |
| COPDGene | 4,000 | 50-90 | 65 | 35 | 0 | 0 | 0 | 0 |
| FHS | 7,574 | 26-108 | 86 | 3 | 4 | 0 | 0 | 7 |
| HCHS/SOL | 8,400 | 30-87 | 0 | 0 | 100 | 0 | 0 | 0 |
| JHS | 2,900 | 38-102 | 0 | 100 | 0 | 0 | 0 | 0 |
| MASALA | 500 | 50-94 | 0 | 0 | 0 | 100 | 0 | 0 |
| MESA | 4,683 | 65-103 | 38 | 27 | 24 | 12 | 0 | 0 |
| NOMAS | 1,267 | 62-106 | 12 | 14 | 72 | 0 | 1 | 0 |
| PrePF | 2,500 | 40-80 | 92 | 3 | 3 | 0 | 0 | 0 |
| REGARDS | 8,000 | 57-105 | 62 | 38 | 0 | 0 | 0 | 0 |
| SARP | 380 | 18-80 | 75 | 25 | 0 | 0 | 0 | 0 |
| SPIROMICS | 1,800 | 47-87 | 82 | 4 | 4 | 0 | 0 | 0 |
| SHS | 2,701 | 31-105 | 0 | 0 | 0 | 0 | 100 | 0 |

NHW = Non-Hispanic White. B = Black. H/L = Hispanic/Latinx. As = Asian American. Am Ind = American Indian. Of note, ARIC did not inquire regarding Hispanic/Latino ethnicity, hence White participants cannot be definitely defined as non-Hispanic.
ARIC = Atherosclerosis Risk in Communities Study; C4R = Collaborative Cohort of Cohorts for COVID-19 Research; CARDIA = Coronary Artery Risk Development in Young Adults; COPDGene= Genetic Epidemiology of COPD; FHS = Framingham Heart Study; HCHS/SOL = Hispanic Community Health Study/Study of Latinos; JHS = Jackson Heart Study; MASALA = Mediators of Atherosclerosis in South Asians Living in America; MESA = Multi-Ethnic Study of Atherosclerosis; NOMAS = Northern Manhattan Study; PrePF = Prevent Pulmonary Fibrosis; REGARDS = REasons for Geographic and Racial Differences in Stroke; SARP = Severe Asthma Research Program; SPIROMICS = Subpopulations and Intermediate Outcome Measures in COPD Study; SHS = Strong Heart Study

The cohorts that comprise C4R have collected detailed data on their participants’ health and behavior for as long as fifty years of follow-up (**Figure 1**). As summarized in **Table 2**, C4R cohorts have performed extensive longitudinal phenotyping of subclinical and clinical disease as well as assessments of laboratory biomarkers, ‘Omics, imaging, diet, behavior, and social determinants of health, and they have extensive biorepositories of stored specimens. Twelve cohorts have geocoding available, supporting participant-level assessment of neighborhood socioeconomic status, exposures to systemic racism, and environmental exposures such as air pollution. All C4R cohorts use similar or identical adjudication protocols to ascertain all-cause mortality. Ten cohorts ascertain cardiovascular events including myocardial infarction, stroke, and heart failure. Eight cohorts ascertain respiratory events such as COPD and asthma exacerbations. Seven cohorts ascertain incident cognitive impairment and/or dementia.

**Figure 1.**
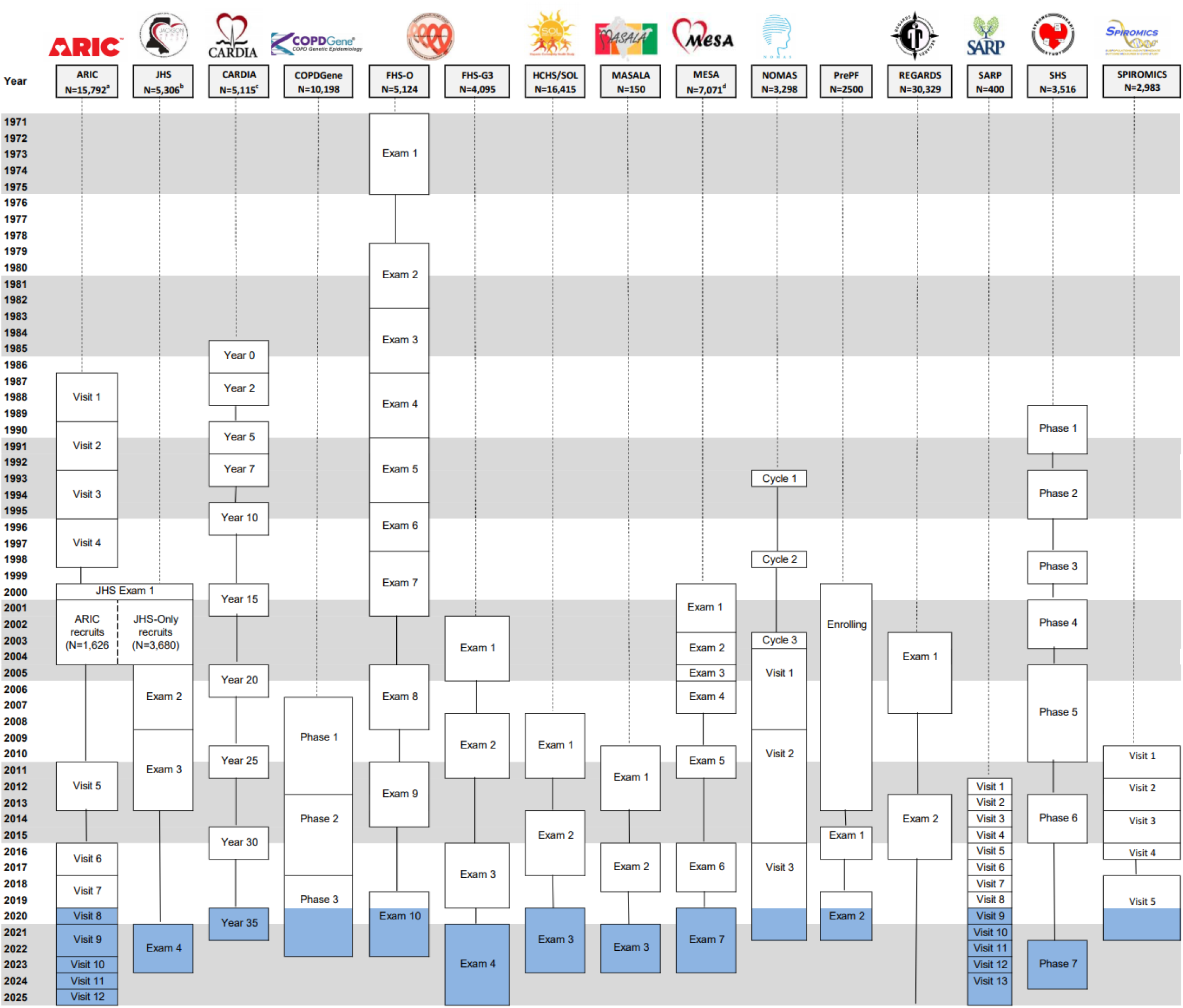
Longitudinal pre-COVID follow up and planned follow-up of C4R participants, by cohort, 1971-2025. Some visits were overlapping, which is not shown; instead, midpoints of the visits are indicated. COVID-era exams are shaded in blue. Solid lines indicate cohort follow up, which typically includes regular contact by telephone and mail and ongoing events ascertainment. ^a^424 gave restricted consent; ^b^ Includes 1,626 participants recruited from ARIC; ^c^ Withdrawal of consent by one participant; ^d^ MESA + 257 new recruits into the MESA Air Pollution Study.

**Table 2.** Density of phenotype data in C4R, 1971-2025.

|  |  | ARIC | CARDIA | COPD Gene | FHS | HCHS/SOL | JHS | MAS-ALA | MESA | NOMAS | PrePF | REG-ARDS | SARP | SPIRO-MICS | SHS |
| --- | --- | --- | --- | --- | --- | --- | --- | --- | --- | --- | --- | --- | --- | --- | --- |
| Incident events/diagnoses | MI, CVA, TIA, HF | A | A | A | A | A | A | A | A | A | A | A | A | A | A |
|  | COPD, asthma | A |  | A |  | A |  |  | A |  | A |  | A | A |  |
|  | Cog. Impairment/Dementia | A | A** |  | A |  | A** |  | A | A |  | A |  |  | A |
|  | Diabetes | A | A |  | A | A | A | A | A | A |  | A |  |  | A |
|  | Chronic kidney disease | A | A |  | A | A | A | A | A | A |  | A |  |  | A |
|  | Mortality | A | A | A | A | A | A | A | A | A | A | A | A | A | A |
| Subclinical disease & Functional measures* | Echocardiography | 2 | 3 |  | 6 | 1 | 1 | 1** | 1 | 1 |  |  |  |  | 3 |
|  | Cardiac MRI |  |  |  | 1 | 1** | 1 |  | 3 |  |  |  |  |  |  |
|  | Carotid IMT/plaque | 4 | 1 |  | 2 |  | 1 | 1 | 3 | 1 |  |  |  |  | 3 |
|  | Brain MRI | 4** | 3** |  | 3 | 1** | 1** |  | 2** | 1 |  |  |  |  | 2 |
|  | CT coronary artery calcium | 1 | 4 | 1 | 2 | 1** | 1 | 2 | 3 | 1 |  |  |  |  |  |
|  | Pulmonary function | 3 | 7** | 3 | 6 | 1 | 1 |  | 3 |  | 1 |  | 6 | 5 | 2 |
|  | CT lung imaging |  | 1** | 3 | 1 |  |  |  | 3 |  | 3 |  | 2 | 3 |  |
|  | Polysomnography | 1 | 2** |  | 1 | 1 | 1 |  | 2* | 1 |  |  |  |  | 2 |
|  | Cognitive assessments | 6* | 3** | 1 | 9 | 3** | 1** |  | 4 | 20 |  | 6* |  | 1 | 2 |
|  | Psychological assessments | 6* | 9** |  | 9 | 3** | 4** | 2** | 7** | 20 |  | 2* |  |  | 6** |
|  | Geocoding | A | A | A | A | A | A | A | A | A |  | A | A | A |  |
| Serologic measures* | Metabolic (lipids, glucose) | 8** | 10** |  | 9 | 3** | 4** | 3** | 7** | 3** |  | 2 |  |  | 6** |
|  | Inflammatory | 8** | 4 |  | 2 | 2** | 4** | 1 | 6** | 2 |  | 2 |  |  | 6** |
|  | Blood counts | 1 |  | 3 | 2 | 3 |  |  | 2 | 2 |  | 2 | 6 | 5 |  |
|  | Renal (blood) | 8** | 10** |  | 6 | 3** | 2** | 3** | 7** | 2 |  | 2 |  |  | 6** |
|  | Renal (urine) | 5** | 2 |  | 7 | 3** | 4** | 1 | 2 |  |  | 2 |  |  | 6** |
|  | Stored blood | 8** | 10** | 3 | 9 | 3** | 4** | 3** | 7** | 2 | 3 | 2 | 6 | 5 | 6** |
| Omics* | GWAS, N | 12527 | 3622 | 10000 | 8482 | 12670 | 3443 |  | 4287 | 1267 | 2500 | 3851 | 1418 | 2900 | 3476 |
|  | WGS (T = TOPMed) | T | T | T | T | T | T |  | T |  | T |  | T | T |  |
|  | RNA Seq-blood | 1 | 2 | 1 | 1 |  | 1 |  | 2 |  |  |  | 2 | 1 |  |
|  | RNA Seq-lung tissue |  |  |  |  |  |  |  |  |  |  |  | 1 | 1 |  |
|  | DNA methylation | 3 | 2 | 2 | 2 |  | 1 | 1 | 2 |  | 1 | 1 |  |  | 1 |
|  | Proteomics | 3 | 1 | 1 | 2 |  | 1 | 1** | 2 |  |  |  |  | 1 |  |
|  | Metabolomics | 2 | 1 |  | 2 | 1 | 1 | 1 | 2 |  |  | 1 |  |  | 2 |

\*Maximum number of measures per participant. \*\*Includes planned measurement at upcoming exam. A = Ascertained. T = on TOPMed. Data reflected in the table may be collected in only a subset of the cohort (e.g., as part of an Ancillary Study).

### Collaboration

Most of the cohorts have a long history of collaboration in the genomics-oriented Cohorts for Heart and Aging Research in Genomic Epidemiology (CHARGE) consortium (39), the NHLBI Pooled Cohorts Study focusing on respiratory epidemiology (40), the Cross-Cohort Collaboration (CCC) for cardiovascular epidemiology (41), the Blood Pressure and Cognition (BP COG) Study (42), and the genetic sequencing and multi-omics-focused Trans-Omics for Precision Medicine (TOPMed) Project (43). C4R is building and expanding upon these successes to advance COVID-19 research.

Planning for C4R began in March 2020, when the need for a coordinated, cross-cohort response to the knowledge gaps posed by the COVID-19 pandemic became self-evident and urgent. Cohort investigators initiated discussions regarding approaches to ascertain SARS-CoV-2 infections and COVID-related illnesses within the context of unprecedented cohort operational challenges associated with the outbreak. The National Heart, Lung, and Blood Institute (NHLBI) funded C4R via an Other Transactional Authority (OTA) mechanism in October 2020. Additional funding for inclusion of the neurology-focused cohorts was provided via the OTA by the National Institutes of Neurological Disorders and Stroke (NINDS) and the National Institute of Aging (NIA).

Leadership for C4R is provided by an organizing committee that includes leading – and often, founding – principal investigators (PIs) from all C4R cohorts, PIs from the C4R Data Coordination and Harmonization Center (DCHC), PIs from the C4R Biorepository and Central Laboratory (BCL), and program officers from the NHLBI, NINDS, and NIA. This organizing committee developed master C4R protocols for COVID-19 data collection.

Consistent with an ancillary studies model, each cohort in C4R is directly responsible for accomplishing its own data collection in accordance with the master protocol and under the supervision of its respective Observational Study Monitoring Board (OSMB), Steering Committee, and any other applicable regulatory authorities.

To promote and sustain this broad collaborative effort, C4R PIs invited additional investigators and cohort personnel to participate in C4R committees and working groups. Study materials, including protocols and meeting materials, are posted regularly on a password-protected investigator section of the C4R website (c4r-nih.org).

### Participants

Cohort participants previously consented for in-person, telephone, and/or email contact and for the abstraction of medical records. Additional consent for ascertainment of COVID-19 data, including the serosurvey, is being obtained according to cohort-specific procedures, including verbal, remote, and traditional written informed consent.

Of 73,119 active participants across the fourteen cohorts, 53,972 participants were readily available for recruitment into C4R. Anticipated socio-demographic characteristics of potential C4R participants, estimated from current active cohort participants, are shown in **Table 3**. Fifty-eight percent of potential participants are 65 years or older, and thus at high risk for severe COVID-19. The anticipated sample is racially and ethnically diverse, based on self-report (44), with approximately 6% American Indian participants, 2% Asian participants, 26% Black participants, and 20% Hispanic/Latino participants.

**Table 3.** Planned enrollment of C4R target population, estimated based on characteristics of current active cohort participants, 2020.

| Cohort | C4R |  |
| --- | --- | --- |
|  | N | % |
| Total | 53,972 | 100.00 |
| Sex |  |  |
| Female | 31,034 | 57.50 |
| Male | 22,938 | 42.50 |
| Race |  |  |
| Asian | 1,255 | 2.33 |
| American Indian and Alaska Native | 3,015 | 5.59 |
| Black | 13,937 | 25.82 |
| Native Hawaiian or Pacific Islander | 32 | 0.06 |
| White | 29,104 | 53.92 |
| Other | 3,156 | 5.85 |
| Multiple | 2,037 | 3.77 |
| Unknown | 1,436 | 2.66 |
| Ethnicity |  |  |
| Hispanic | 10,862 | 20.12 |
| Non-Hispanic | 34,899 | 64.66 |
| Unknown | 8,211 | 15.21 |
| Age group |  |  |
| 18-29 | 663 | 1.23 |
| 30-64 | 21,920 | 40.61 |
| 65+ | 31,389 | 58.16 |

All forty-eight continental states are represented among C4R participants, including rural, suburban, and urban communities (**Figure 2**). In all, C4R is being conducted across forty field/clinical centers, many of which are associated with more than one C4R cohort; one cohort with extensive geographic reach, the REGARDS, operates via telephone and in-home exams only (38).

**Figure 2.**
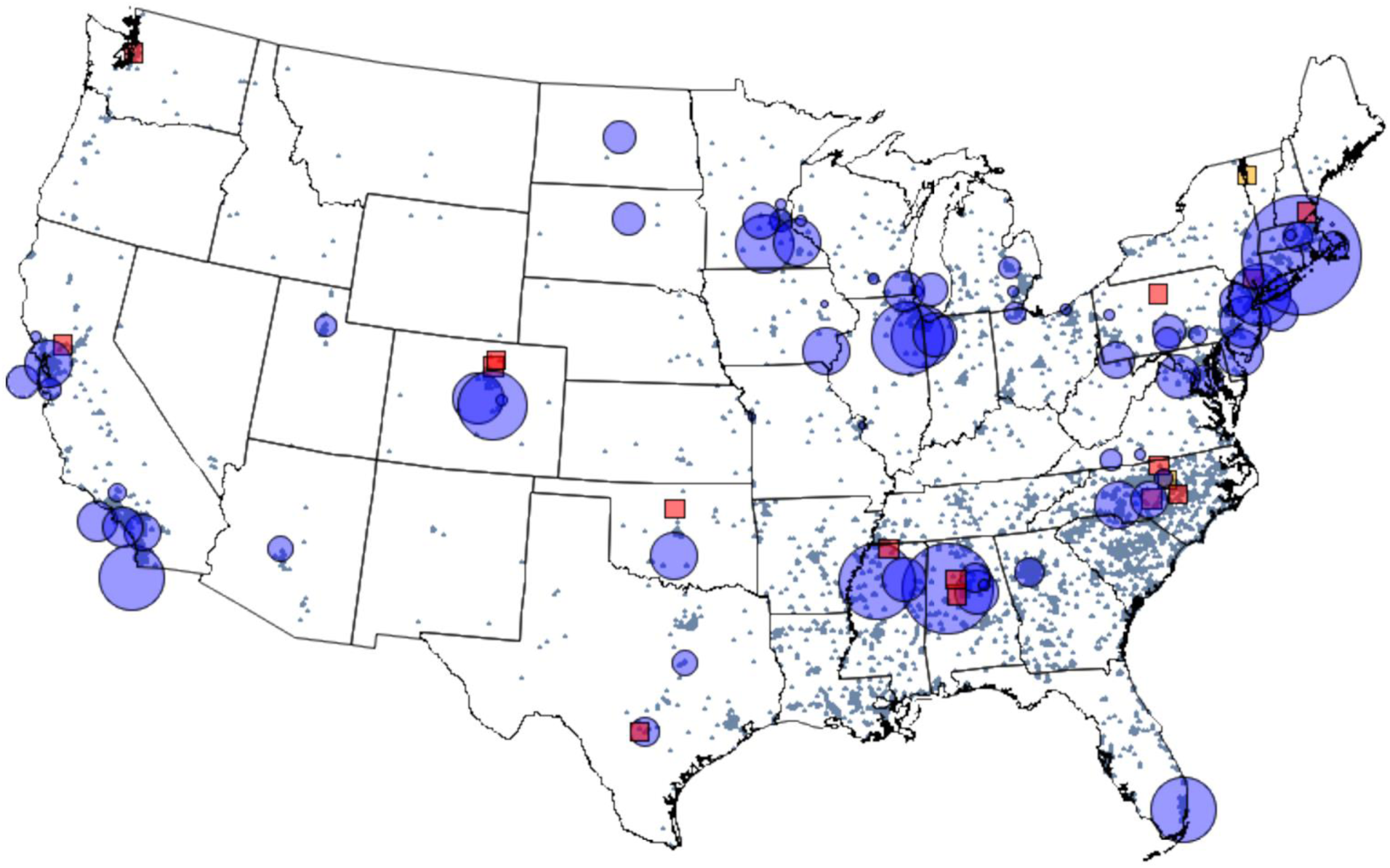
C4R participants, field/clinical centers, and coordinating centers. Blue circles indicate field/clinical centers, and the size is proportional to the number of participants at that field/clinical center. Participants in the REGARDS, which does not have field/clinical centers, are shown by additional blue shading according to their geocoded home addresses. Red squares indicate coordinating centers involved in the study. Yellow squares indicate C4R central resources: the data coordination and harmonization center, the biorepository and central laboratory, and the administrative coordinating center.

### Data collection

#### COVID-19 questionnaires

C4R is ascertaining self-reported COVID-related experiences by questionnaire. Each cohort will deploy C4R questionnaires twice within 18 months following the initial outbreak in March 2020 via telephone, mail-in, online, email, or smartphone apps. Wave 1 questionnaires were developed as early as March 2020 in certain cohorts (45) and urgently administered in spring and summer 2020. Although these efforts pre-dated C4R, early informal cross-cohort collaborations ensured that many cohorts used identical questionnaires, and all of them generated common data elements regarding infection, testing, hospitalization, and recovery. Wave 2 questionnaires were fully standardized to include domains on COVID-19 infection, testing, hospitalization, symptoms, recovery, re-infection, contacts, vaccination, behavioral changes, sleep, memory loss, depression, anxiety, fatigue, and resilience. The C4R questionnaire was developed collaboratively to include validated and PhenX toolkit instruments (https://www.phenxtoolkit.org) (46-55) in order to optimize comparability with pre-pandemic assessments and across C4R and other epidemiology cohorts. The C4R questionnaire, including translations into Spanish and Mandarin, are available on PhenX; Research Electronic Data Capture (REDCap (56, 2)) programming may be available on request.

#### COVID-related events ascertainment

C4R is ascertaining COVID-related hospitalizations and deaths that are identified via the C4R questionnaire or other surveillance methods available to the cohorts, including EHR linkages, where available. Each cohort is using its own established infrastructure for ascertainment of medical records and death certificates, including use of the National Death Index (NDI), the Centers for Medicare & Medicaid Services (CMS), International Classification of Diseases (ICD) codes (58), and linkage to records from local departments of health. Cohorts may review events locally at their Field/Coordinating Centers or transfer records for central review by C4R. The C4R events review is designed to assess severity and major complications of COVID-19 illness, including pneumonia, myocardial infarction, stroke, thromboembolism, and acute renal failure. The protocols use, or are modeled after, longstanding cohort protocols to classify and validate cardiovascular, respiratory (19), and thromboembolic (59) events. Protocols for ascertainment, review, and classification are available on the study website (<u>c4r-nih.org</u>).

#### Dried blood spot collection

C4R is ascertaining serostatus by dried blood spot (DBS) in 2021. Cohort field centers receive DBS collection kits from the BCL and are responsible for recruitment, consent, and distribution to participants. Updated details regarding vaccination status are obtained at the time of DBS consent and immediately prior to mailing the DBS kit to the participant. Participants mail the completed kits directly to the BCL or to the cohort field or coordinating center as an intermediary step. Participant instructions, including a video, are provided by the cohort and via the C4R website (<u>c4r-nih.org</u>) and/or cohort-specific websites. In cohorts with upcoming in-person exams, the DBS may be collected in-person by research staff.

### C4R Common Data Elements

C4R data collection will define a spectrum of COVID-19 outcomes. Ascertainment of COVID-related hospitalizations and deaths will characterize, classify, and validate moderate-to-severe COVID-19 illnesses. In addition to identifying these events, questionnaires are being used to obtain self-reported information on the nature, severity, and duration of symptoms during acute infection and in the post-acute setting. This will support classification of symptomatic and asymptomatic infections, as well as cases of prolonged recovery or post-acute sequelae of SARS-CoV-2 infection (PASC). Data on behaviors, attitudes, psychosocial impacts, and vaccinations will also be collected. Seropositive individuals without self-reported infection will be reclassified as infected, whereas seronegative individuals with prior positive testing by self-report or health records will be classified as sero-reverted.

### Data management

C4R data collection is coordinated centrally at the DCHC at Columbia University Irving Medical Center. Electronic data collection forms are being programmed into REDCap for use or adaptation by the cohort coordinating centers. Metadata on completion of questionnaires, events ascertainment, and DBS kit collection status are reported and reviewed bi-weekly to ensure operational milestones are met. Participants are assigned a C4R study identifier by cohort-specific coordinating centers that is used for participant-level data transfers and analyses.

### Biorepository and central laboratory and serology measurement

The C4R BCL at the University of Vermont is responsible for establishing a C4R biorepository of DBS, plus other biospecimens that may be collected in the future, and for performing and/or coordinating performance of any centralized clinical and biomarker assays and serology assays. Individual DBS Collection kits are produced by the BCL and shipped to the cohorts (either to the individual field centers or the cohort coordinating center, based on cohort preference). Kits and DBS cards are labeled with a biospecimen identifier, which is linked to C4R identifiers that are maintained centrally and not shared with the BCL, through the use of a “linking key.” Filled DBS cards are returned to the BCL, and batches prepared for serology assays performed by the New York State Wadsworth Center’s Bloodborne Viruses Laboratory (BVL) under CLIA and New York State certification. The BVL performs a SARS-CoV-2 IgG Microsphere Immunoassay using Luminex bead technology for qualitative detection of human IgG antibodies to SARS-CoV-2 nucleocapsid (N) and spike subunit 1 (S1) antigens. Based on testing 730 pre-COVID DBS and >1100 DBS from individuals with laboratory-confirmed infection, specificity is 99.5% for both N and S1 and sensitivity ranged from 90 to 96% for symptomatic individuals and 77 to 91% for asymptomatic individuals. Sensitivity increased for both groups with time from positive PCR test, accounting for the range. This assay was used successfully to test over 57,000 DBS for statewide serosurveys from April-June as part of New York State’s public health response. Serology results are reported by the BVL to the C4R BCL, and then to the cohort coordinating centers, which are responsible for a) recombining the results with the proper participants based on the “linking key”, and b) reporting results to participants according to usual cohort practices. Serological results are not believed to have clinical relevance, and the CDC does not currently recommend modifications to individual behavior or clinical care based on antibody status alone (60); hence, no protocols for “alert” findings have been established, and participants may opt out of results return. Protocols for the serosurvey are available on the study website (<u>c4r-nih.org</u>).

Since all current vaccines in use in the U.S. generate an immune response to the Spike protein, we anticipate being able to distinguish vaccination from viral infection by the use of the anti-nucleocapsid assay results (61).

### Harmonization

Harmonization of COVID-19 and pre-pandemic data will be performed centrally to define COVID-19 common data elements and to align pre-pandemic data for large-scale, longitudinal analyses. This effort will leverage prior harmonization efforts across C4R cohorts in the TOPMed Project, the NHLBI Pooled Cohorts Study, the BP COG Study, and the CHARGE Working Groups (10, 40, 42, 62-67). Due to their significance to COVID-19 epidemiology, particular emphasis will be placed on harmonizing the large amount of deep pre-pandemic physiologic (40), neurocognitive (68-74), and imaging-based (75-82) phenotyping collected within the decade prior to the outbreak using deep-learning (18, 83-85) and other methods (**Table 4**).

**Table 4.**
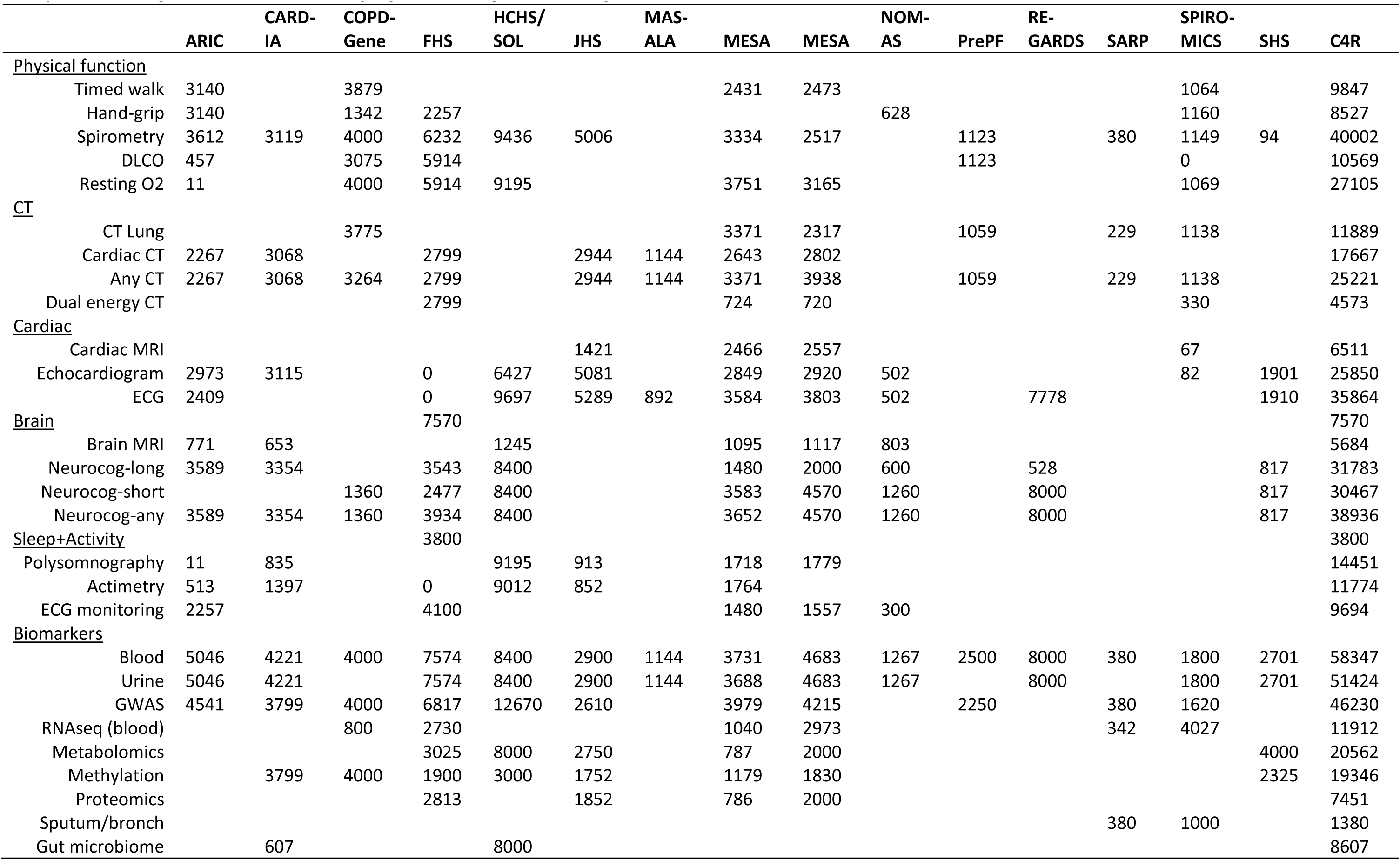
Estimated number of participants with recent pre-pandemic deep phenotyping for harmonization in C4R, by cohort, 2010-2020. If the most recent exam was prior to 2010, data are not included. CT = computed tomography. ECG = electrocardiogram. GWAS = genome-wide association study. MRI = magnetic resonance imaging. Neurocog = neurocognitive.

### Quality control

C4R cohorts have established protocols for checking data completeness and accuracy at the field center and coordinating center levels. Dual data entry is encouraged but not required, since it will not be feasible in all settings due to local impediments and COVID-related exigencies. Ten percent of event reviews will be randomly selected for re-review. Reviewers not meeting standards will receive regular feedback with recommendations for retraining and/or protocol modifications, as appropriate. Serological assays will be repeated on a random 5% sub- sample of blind duplicates.

### Data sharing

The C4R Commons Agreement, modeled on the CHARGE Analysis Commons Consortium Agreement (86), will expedite cross-cohort data harmonization and sharing, as allowed (87). Following review and approval, cohort-specific agreements would permit COVID-19 and pre-pandemic data to be uploaded to the NIH-supported cloud computing platform, hosted by BioData Catalyst. Access to the pooled C4R dataset would be granted to investigators involved in core harmonization efforts and those with manuscript proposals approved by C4R publications and cohort coordinating committees. Once harmonization and related quality control is completed, C4R common data elements will be transferred as a limited dataset for public access on BioData Catalyst in accord with cohort-specific consents and commitments.

### Governance

The administrative coordinating center for C4R is the NHBLI CONNECTS program (nhlbi- connects.org). Metadata on operational progress is submitted biweekly to CONNECTS for tracking and review purposes. Central functions of C4R are overseen by a C4R OSMB convened by CONNECTS.

## DISCUSSION

C4R will leverage existing American cohort studies to develop a large, multi-ethnic, pooled cohort of participants with incident COVID-19 and COVID-free participants that is relatively free of referral, survival, and recall biases compared to clinically based inception cohorts of COVID- 19 patients. C4R includes a highly diverse population of US adults, including older and socially disadvantaged populations that have especially high risk of adverse COVID-19 outcomes. C4R is distinguished from other large studies of COVID-19 by its unparalleled wealth of pre-pandemic phenotyping, providing unique opportunities to evaluate a range of risk and resilience factors for SARS-CoV-2 infection and adverse COVID-19 outcomes, including severe COVID-19 illness, PASC, and other long-term effects of the pandemic response. Unlike case registries and EHR-based studies, C4R’s repeated exams and cognitive assessments before and after COVID-19 also provide important opportunities to estimate the social and behavioral impact of the COVID-19- related pandemic response on changes in long-term mental and physical health across multiple domains.

C4R will provide important opportunities for future studies using a range of epidemiologic study designs. For example, nested within C4R, longitudinal cohort studies of COVID-affected and unaffected participants could repeat a variety of subclinical measures (e.g., echocardiography, lung imaging, neuro-cognitive assessment) to define reliably the consequences of COVID-19 infection. Ongoing high-quality events follow-up will allow assessment of long-term clinical health outcomes following COVID-19 and the pandemic period. The extensive biobanks maintained by the cohorts could support measurement of prior viral infections, immune-phenotypes, metabo-types, ‘Omics, and other pre-COVID characteristics that may be risk determinants or modifiers for COVID-19 susceptibility and vaccine effectiveness. The fact that the cohorts continue to follow their participants provides a dynamic resource to study emerging questions in COVID-19 epidemiology, including but not limited to viral variants and vaccination. And, C4R provides a model for cross-cohort collaboration and active data sharing that will promote consortium-based epidemiologic work on biological, social, and epidemiologic questions beyond the COVID-19 pandemic, in alignment with recommendations for the strategic transformation of population studies (88).

## Data Availability

Following review and approval, cohort-specific agreements would permit COVID-19 and pre-pandemic data from C4R to be uploaded to the NIH-supported cloud computing platform, hosted by BioData Catalyst.

**Author affiliations**

**Albert Einstein College of Medicine**

- Department of Epidemiology and Population Health, Albert Einstein College of Medicine, Bronx, New York, United States (Carmen R Isasi, Robert C Kaplan)
- Children’s Hospital/ Harvard Medical School

**Boston Children’s Hospital/Harvard Medical School**

- Department of Medicine, Division of Allergy and Immunology, Boston Children’s Hospital, Boston Massachusetts (Wanda Phipatanakul)

**Boston University**

- Department of Pathology & Laboratory Medicine, Boston University School of Medicine, Boston, Massachusetts, United States (Joel M Henderson)
- Departments of Medicine and Epidemiology, Boston University, Boston, Massachusetts, United States (Ramachandran S Vasan)
- Department of Medicine, School of Medicine, and Department of Biostatistics, School of Public Health, Boston University, Boston, Massachusetts, United States (Vanessa Xanthakis)

**Brigham and Women’s Hospital/ Harvard Medical School**

- Pulmonary and Critical Care Medicine, Brigham and Women’s Hospital, Boston, Massachusetts (Bruce Levy, Matthew R Moll)
- Channing Division of Network Medicine, Brigham and Women’s Hospital, Boston, MA (Matthew Moll).

**Colorado Anschutz Medical Campus**

- Division of Pulmonary Sciences and Critical Care, Department of Medicine, University of Colorado Anschutz Medical Campus, Aurora, Colorado, United States (Alyssa Asaro, Joyce Lee, David Schwartz)
- Department of Epidemiology, School of Public Health, University of Colorado Anschutz Medical Campus, Aurora, Colorado, United States (Gregory Kinney)

**Columbia University**

- Division of General Medicine, Department of Medicine, Columbia University Medical Center, New York, New York, United States (Pallavi P Balte, R Graham Barr, Akshaya Krishnaswamy, Elizabeth C Oelsner, Priya Palta, Yiyi Zhang)
- Columbia Data Coordinating Center, New York State Psychiatric Institute and Columbia College of Physicians and Surgeons, Columbia University Medical Center, New York, New York, United States (Howard F Andrews)
- Department of Neurology, Vagelos College of Physicians and Surgeons, Columbia University, New York, New York, United States (Mitchell Elkind, Jose Gutierrez, Jennifer J Manly)
- Department of Epidemiology, Mailman School of Public Health, New York, NY United States (Ryan T Demmer [adjunct], Mitchell Elkind)

**Georgetown University**

- MedStar Health Research Institute and Department of Medicine, Georgetown University, Washington, District of Columbia, United States (Jason G Umans)

**Johns Hopkins University**

- Department of Epidemiology, Bloomberg School of Public Health, Johns Hopkins University, Baltimore, Maryland, United States (Josef Coresh)
- Division of Cardiology, Department of Medicine, Johns Hopkins University, Baltimore, Maryland, United States (Wendy S Post)

**Lundquist Institute**

- Lundquist Institute, Los Angeles, California, United States (Jerome I Rotter)

**National Institutes of Health (intramural)**

- Division of Intramural Research, National Heart Lung and Blood Institute of the National Institutes of Health (Veronique Roger)

**National Jewish Health**

- Division of Pulmonary, Critical Care & Sleep Medicine, Department of Medicine, National Jewish Health, Denver, Colorado, United States (Barry Make)
- Division of Rheumatology, Department of Medicine, National Jewish Health, Denver, Colorado, United States (Elizabeth A Regan)
- National Jewish Health, Denver, Colorado, United States (Grace Chen)

**New York State Department of Health**

- Bloodborne Viruses Laboratory, Wadsworth Center, New York State Department of Health, Albany, New York, United States (Monica Parker)

**Northwestern**

- Center for Epidemiology and Population Health, Department of Preventive Medicine, Northwestern University, Chicago, Illinois, United States (Norinna Bai Allen, Namratha R Kandula)
- Division of General Internal Medicine, Department of Medicine, Northwestern University, Chicago, Illinois, United States (Namratha R Kandula)

**Pennsylvania State University**

- Division of Biostatistics and Bioinformatics, Department of Public Health Sciences, Pennsylvania State University, State College, Pennsylvania, United States (Dave Mauger)

**San Diego State University**

- South Bay Latino Research Center, Department of Psychology, San Diego State University, San Diego, California, United States (Linda Gallo, Gregory A Talavera)

**Texas Biomedical Research Institute**

- Population Health Program, Texas Biomedical Research Institute, San Antonio, Texas, United States (Shelley A Cole)

**University of Alabama at Birmingham**

- Department of Biostatistics, University of Alabama at Birmingham, Birmingham, Alabama, United States (Suzanne E Judd)
- Department of Epidemiology, University of Alabama at Birmingham, Birmingham, Alabama, United States (Kelley Pettee Gabriel, Virginia J Howard, Cora E Lewis, Emily B Levitan)
- Division of Preventive Medicine, School of Medicine, University of Alabama at Birmingham, Birmingham, Alabama, United States (James M Shikany)

**UCLA**

- The Institute for Translational Genomics and Population Sciences, Department of Pediatrics, The Lundquist Institute for Biomedical Innovation at Harbor-UCLA Medical Center, Torrance, CA USA (Jerome I Rotter)
- Division of Cardiology, Department of Medicine, UCLA Medical Center, Los Angeles, California, United States (Karol E Watson)

**University of California San Francisco**

- Department of Medicine, University of California San Francisco, San Francisco, California, United States (Alka M. Kanaya, Arunee A. Chang)
- Department of Obstetrics, Gynecology & Reproductive Sciences, University of California San Francisco, San Francisco, California, United States (Michael Schembri)
- Division of Pulmonary, Critical Care, Allergy, and Sleep Medicine, Department of Medicine, University of California San Francisco, San Francisco, California, United States (Prescott G. Woodruff)

**University of Kentucky**

- Department of Epidemiology, College of Public Health, University of Kentucky, Lexington, KY (Anna M Kucharska-Newton).

**University of Miami**

- Department of Neurology and Evelyn F. McKnight Brain Institute, University of Miami, Miami, Florida, United States (Tatjana Rundek, Ralph L. Sacco)

**University of Michigan**

- Division of Pulmonary and Critical Care, Department of Medicine, University of Michigan, Ann Arbor, Michigan, United States (MeiLan K. Han)
- Division of General Medicine, Department of Medicine, University of Michigan, Ann Arbor, Michigan, United States (Deborah A Levine)

**University of Minnesota**

- Division of Epidemiology and Community Health, School of Public Health, University of Minnesota, Minneapolis, Minnesota, United States (Ryan T Demmer, Aaron R Folsom, David R Jacobs Jr)

**University of Mississippi Medical Center**

- Departments of Medicine and Population Health Science, School of Public Health, University of Mississippi Medical Center, Jackson, Mississippi, United States (Pramod Anugu, Adolfo Correa, Mario Sims, Yuan-I Min)
- School of Nursing, University of Mississippi Medical Center, Jackson, Mississippi, United States (Karen Winters)

**University of North Carolina**

- Collaborative Studies Coordinating Center, Department of Biostatistics, Gillings School of Global Public Health, University of North Carolina, Chapel Hill, North Carolina, United States (David Couper, Kimberly Ring)
- Department of Epidemiology, Gillings School of Global Public Health, University of North Carolina, Chapel Hill, North Carolina, United States (Anna Kucharska-Newton)
- Department of Nutrition, Gillings School of Global Public Health, University of North Carolina, Chapel Hill, North Carolina, United States (Katie Meyer)

**University of Oklahoma Health Sciences Center**

- Center for American Indian Health Research, Department of Biostatistics and Epidemiology, Hudson College of Public Health, University of Oklahoma Health Sciences Center, Oklahoma City, Oklahoma, United States (Tauqeer Ali, Kimberly Malloy, Ying Zhang)

**University of Pittsburgh**

- Department of Environmental and Occupational Health, Graduate School of Public Heath, University of Pittsburgh, Pittsburgh, Pennsylvania, United States (Sally E Wenzel)
- Division of Pulmonary, Allergy, and Critical Care Medicine, Department of Medicine, University of Pittsburgh, Pittsburgh, Pennsylvania, United States (Jessica Bon)

**UT Health San Antonio**

- Glenn Biggs Institute for Alzheimer’s & Neurodegenerative Diseases, Graduate School of Biomedical Sciences, UT Health San Antonio, San Antonio, Texas, United States (Sudha Seshadri)

**University of Vermont**

- Laboratory for Clinical Biochemistry Research, Department of Pathology & Laboratory Medicine, Larner College of Medicine, University of Vermont, Burlington, Vermont, United States (Rebekah Boyle, Elaine Cornell, Russell Tracy)
- Department of Medicine, Vermont Center for Cardiovascular and Brain Health, Larner College of Medicine at the University of Vermont, Burlington, Vermont, United States (Mary Cushman, Debora Kamin Mukaz)

**University of Washington**

- Department of Epidemiology, School of Public Health, University of Washington, Seattle, Washington, United States (Amanda M Fretts, Robert Kaplan, Bruce Psaty)
- Department of Health Services, School of Public Health, University of Washington, Seattle, Washington, United States (Bruce M Psaty)
- Division of General Medicine, Department of Medicine, University of Washington, Seattle, Washington, United States (Bruce Psaty)
- Department of Biostatistics, School of Public Health, University of Washington, Seattle, Washington, United States (Karen Hinckley Stukovsky)

**Wake Forest School of Medicine**

- Department of Epidemiology and Prevention, Department of Medicine, Wake Forest School of Medicine, Winston-Salem, North Carolina, United States (Alain G Bertoni)
- Division of Pulmonary, Critical Care, Allergy, and Immunologic Diseases, Department of Medicine, Wake Forest School of Medicine (Wendy C Moore, Victor E Ortega)

## Author contributions

All authors meet ICJME criteria for authorship.

## Funding

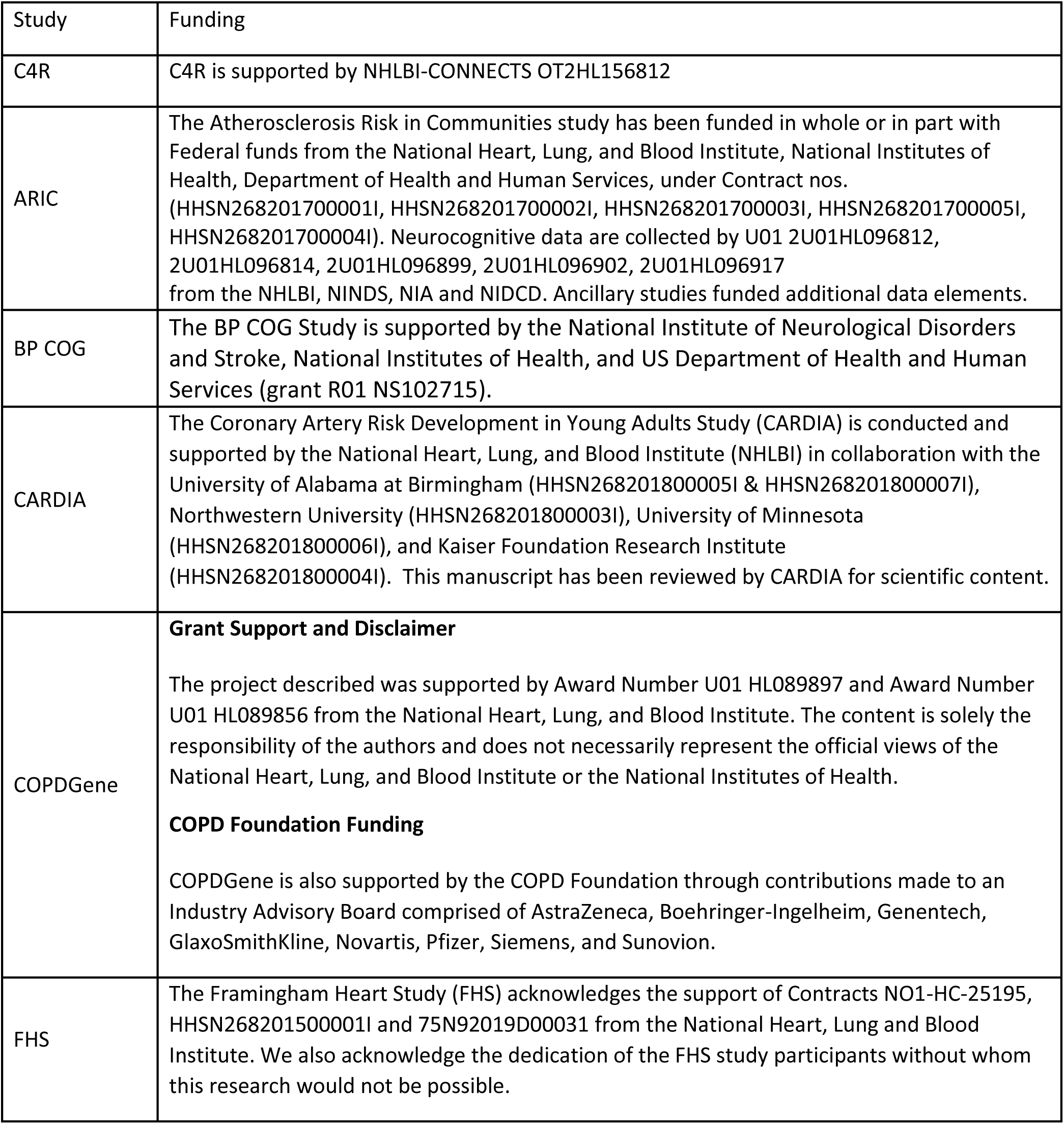
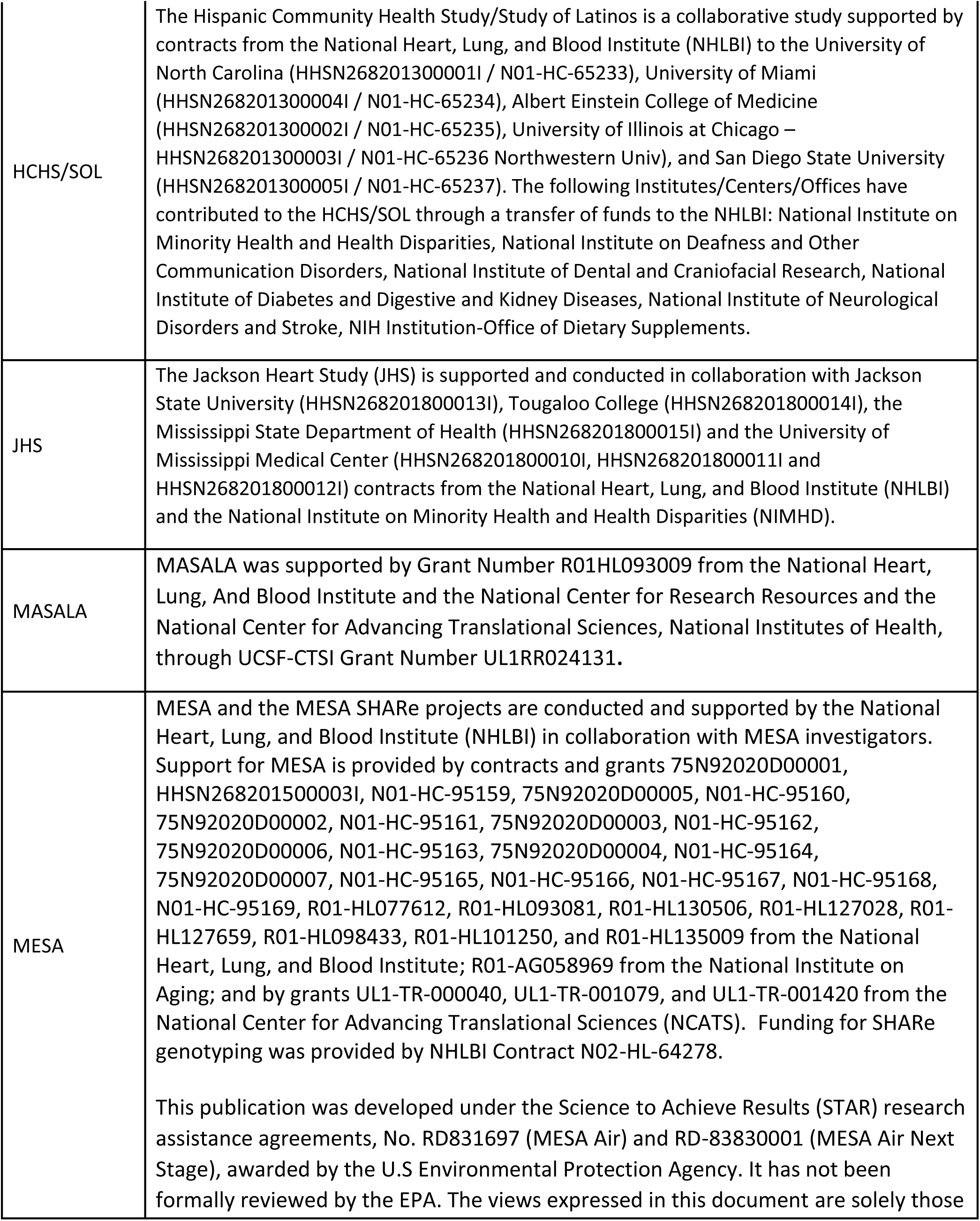
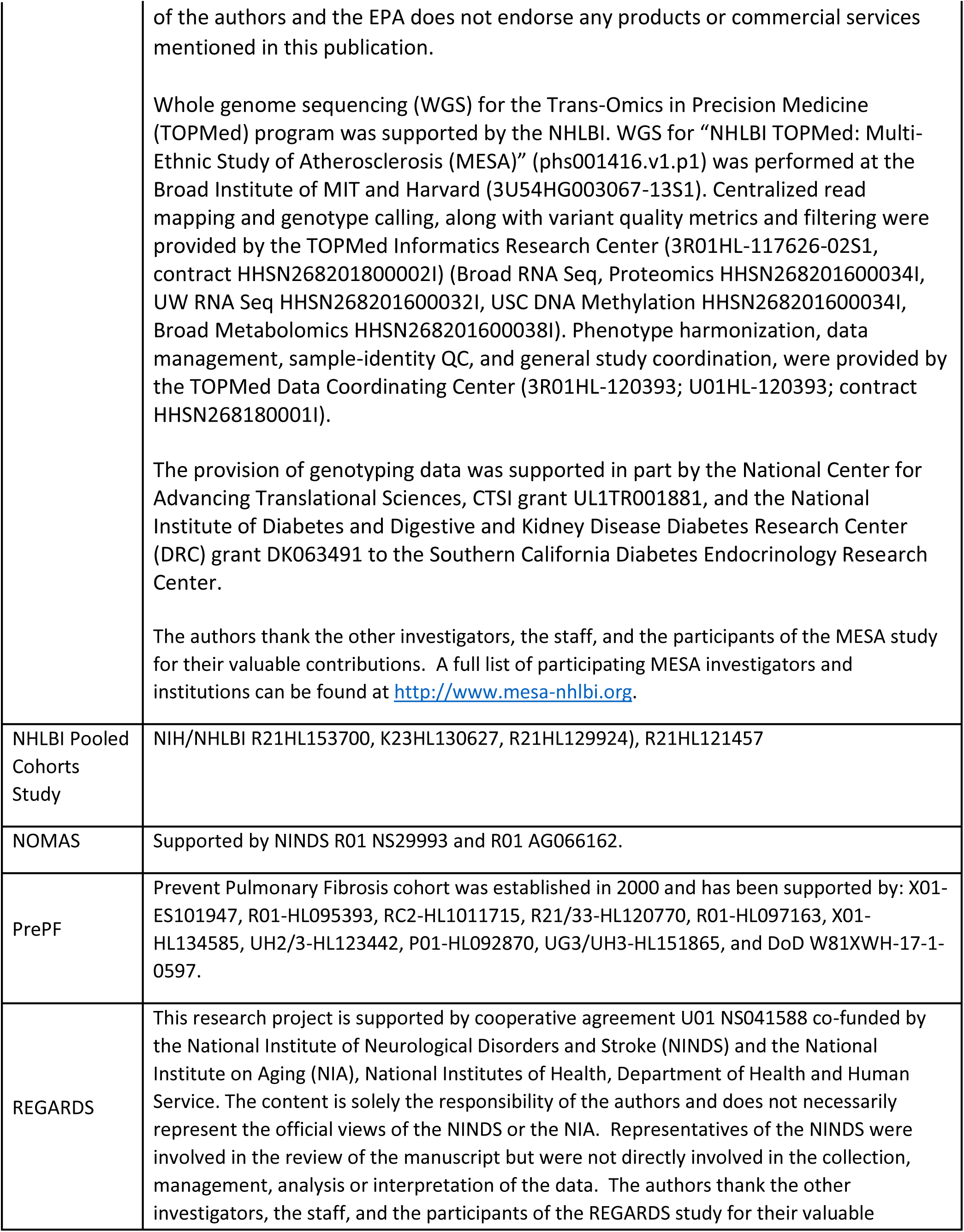
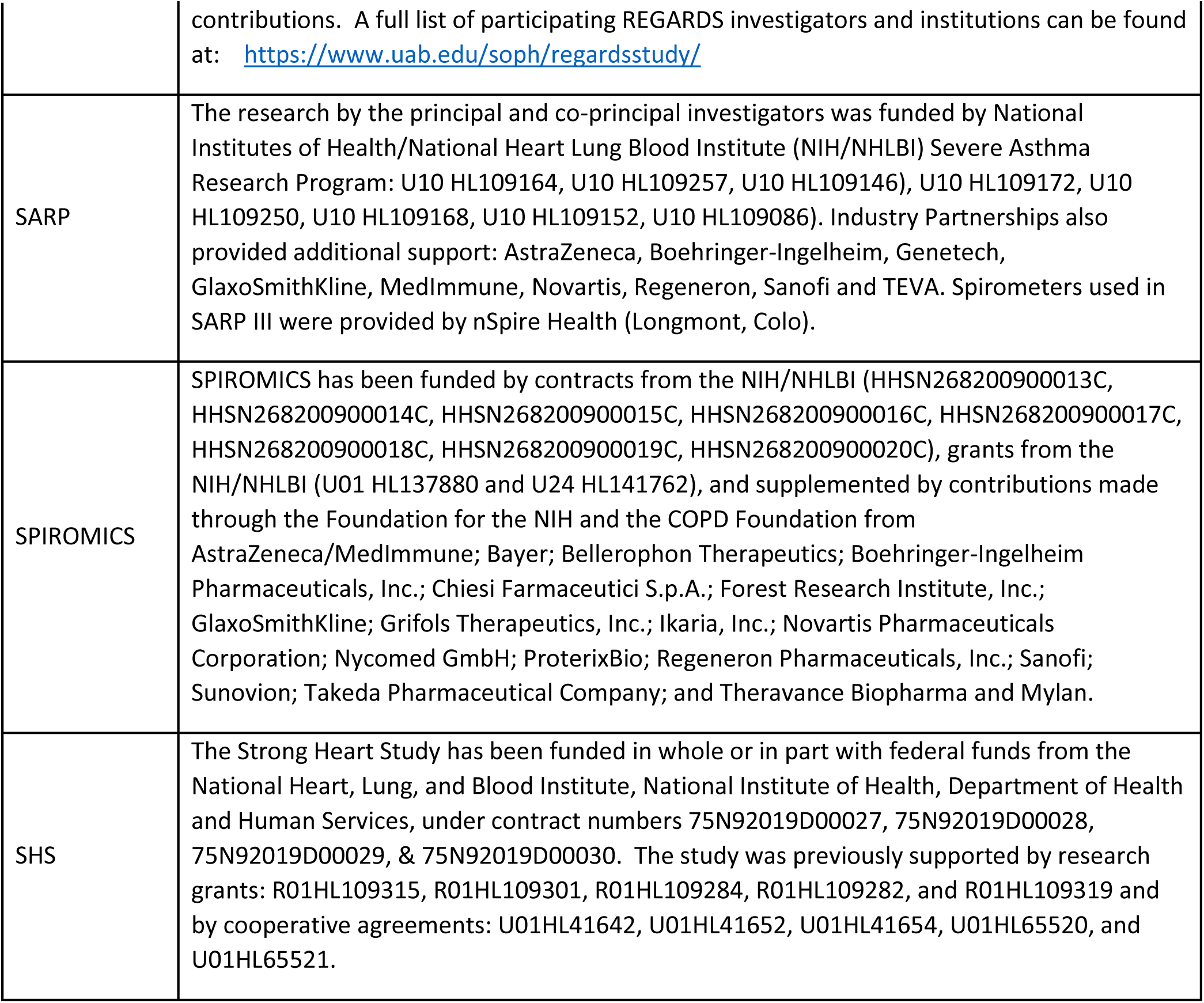

## Thank you’s

We thank the participants of each cohort for their dedication to the studies.

## Members of the study group

Ali, Tauqueer

Allen, Nori

Andrews, Howard

Anugu, Pramod

Arora, Komal

Arynchyn, Alex

Asaro, Alyssa

Balte, Pallavi P

Bancks, Mike

Barr, R Graham

Bateman, Lori

Bertoni, Alain

Bick, Alexander G

Bleecker, Eugene

Bluemke, David

Boyle, Rebekah

Budoff, Matt

Cardwell, Jonathan

Carr, Jeffrey

Causey, Jackie

Chang, Ann

Chen, Grace

Coady, Sean

Cole, Shelley

Coresh, Josef

Cornell, Elaine

Correa, Adolfo

Couper, David

Culbertson, Emily

Curtis, Jeffrey L

Cushman, Mary

DeCarli, Charles

Demmer, Ryan T

Devereux, Richard B

DiPersio, Nicholas

Dolezal, Brett

Doyle, Margaret

Elkind, Mitch

Fain, Sean

Feinstein, Matt

Floyd, James

Freeman, Christine

Fretts, Amanda

Gabriel, Kelley

Gallo, Linda

Gharib, Sina

Goode, Caroline

Goyal, Parag

Griswold, Mike

Han, MeiLan

Hanna, David

Hastie, Annette

Heckbert, Susan R.

Henderson, Joel

Hinckley Stukovsky, Karen

Hoffman, Eric

Hoffman, Udo

Hornig, Mady

Howard, Virginia J

Isasi, Carmen R

Jacobs, David R Jr

Jaquis, Cashell

Judd, Suzanne E

Kamin Mukaz, Debora

Kanaya, Alka M

Kandula, Namratha R

Kaplan, Robert

Khan, Sadiya

King, Jonathan

Kinney, Gregory L

Kucharska-Newton, Anna

Kumi, Smith

Laine, Andrew F.

Lee, Joyce

Lemaitre, Rozenn

Levin, Bonnie

Levine, Deborah

Levitan, Emily B.

Levy, Bruce

Lima, Joao

Lynch, David Make, Barry

Malloy, Kimberly

Manly, Jennifer J

Mauger, Dave

Melius, Katy

Mendoza-Puccini, Carolina

Merkin, Sharon

Meyer, Katie

Meyers, Deborah

Min, Yuan-I (Nancy)

Minotti, Melissa

Moise, Nathalie

Moore, Wendy

Moy, Claudia

Mutalik, Karen

Nasrallah, Ilya

Nelson, Cheryl

Nelson, Lauren

Noel, Patricia

Nordvig, Anna

O’Connor, George

Odden, Michelle

Oelsner, Elizabeth

O’Leary, Marcia

O’Neal, Wanda

Ortega, Victor

Palta, Priya

Pamir, Nathalie

Papanicolaou, George

Phillips, Brenda

Phipatanakul, Wanda

Plante, Tim

Pokharel, Yashashwi

Post, Wendy

Postow, Lisa

Psaty, Bruce

Purcell, Shaun M.

Raffield, Laura

Ramachandran, Vasan

Redline, Susan

Regan, Liz

Reiner, Alex

Rodriguez, Carlos

Roger, Veronique

Rotter, Jerry

Sacco, Ralph

Safford, Monika M.

Schembri, Michael

Schwartz, David

Seshadri, Sudha

Shah, Amil

Shah, Sanjiv J

Shikany, James

Sims, Mario

Smith, Kumi

Smoller, Sylvia

Soliman, Elsayed

Solomon, Scott

Sotres-Alvarez, Daniela

Stewart, Meg

Strobino, Kevin

Sutherland, Patrice

Swenor, Bonnie

Talavera, Gregory

Terry, Greg

Tracy, Russ

Tse, Janis

Umans, Jason

Vasan, Ramachandran S

Vuga, Louis

Wagster, Molly

Wang, Henry

Washko, George

Wentzel, Sally

West, Cynthia

Wilson, Carla

Woodruff, Prescott

Wright, Jackie

Xanthakis, Vanessa

Yuan, Ya

Zakai, Neil

Zhang, Ying

Zhang, Yiyi

## Disclaimer

The content is solely the responsibility of the authors and does not necessarily represent the official views of the National Institutes of Health.

## Abbreviations

ARIC: Atherosclerosis Risk in Communities
BCL: Biorepository and Central Laboratory
C4R: Collaborative Cohort of Cohorts for COVID-19 Research
CARDIA: Coronary Artery Risk Development in Young Adults
CONNECTS: Collaborating Network of Networks for Evaluating COVID-19 and Therapeutic Strategies
COPDGene: Genetic Epidemiology of COPD
FHS: Framingham Heart Study
HCHS/SOL: Hispanic Community Health Study/Study of Latinos
JHS: Jackson Heart Study
MASALA: Mediators of Atherosclerosis in South Asians Living in America
MESA: Multi-Ethnic Study of Atherosclerosis
NOMAS: Northern Manhattan Study
PASC: Post-Acute Sequelae of SARS-CoV-2 Infection
PrePF: Prevent Pulmonary Fibrosis
REGARDS: REasons for Geographic and Racial Differences in Stroke
SARP: Severe Asthma Research Program
SPIROMICS: Subpopulations and Intermediate Outcome Measures in COPD Study
SHS: Strong Heart Study

## References

1. Kissler SM, Tedijanto C, Goldstein E, Grad YH, Lipsitch M. Projecting the transmission dynamics of SARS-CoV-2 through the postpandemic period. Science. 2020.

2. CDC COVID Data Tracker 2021 [Available from: https://covid.cdc.gov/covid-data-tracker/#cases_casesper100klast7days.

3. COVIDview. A weekly surveillance summary of US COVID-19 activity: Centers for Disease Control and Prevention; 2021 [Available from: https://www.cdc.gov/coronavirus/2019-ncov/covid-data/covidview/index.html.

4. Woolf SH, Chapman DA, Lee JH. COVID-19 as the Leading Cause of Death in the United States. JAMA. 2021;325(2):123–4.

5. Andrasfay T, Goldman N. Reductions in 2020 US life expectancy due to COVID-19 and the disproportionate impact on the Black and Latino populations. Proc Natl Acad Sci U S A. 2021;118(5).

6. Huang C, Huang L, Wang Y, Li X, Ren L, Gu X, et al. 6-month consequences of COVID-19 in patients discharged from hospital: a cohort study. The Lancet. 2021;397(10270):220–32.

7. Haynes N, Cooper LA, Albert MA, Association of Black Cardiologists. At the Heart of the Matter: Unmasking and Addressing COVID-19’s Toll on Diverse Populations. Circulation. 2020.

8. Chowkwanyun M, Reed AL, Jr. Racial Health Disparities and Covid-19 - Caution and Context. N Engl J Med. 2020.

9. Rodriguez F, Solomon N, de Lemos JA, Das SR, Morrow DA, Bradley SM, et al. Racial and Ethnic Differences in Presentation and Outcomes for Patients Hospitalized with COVID-19: Findings from the American Heart Association’s COVID-19 Cardiovascular Disease Registry. Circulation. 2020.

10. Oelsner EC, Balte PP, Grams ME, Cassano PA, Jacobs DR, Barr RG, et al. Albuminuria, Lung Function Decline, and Risk of Incident Chronic Obstructive Pulmonary Disease. The NHLBI Pooled Cohorts Study. Am J Respir Crit Care Med. 2019;199(3):321–32.

11. Livingston E, Bucher K. Coronavirus Disease 2019 (COVID-19) in Italy. JAMA. 2020.

12. Lederlin M, Bauman G, Eichinger M, Dinkel J, Brault M, Biederer J, et al. Functional MRI using Fourier decomposition of lung signal: reproducibility of ventilation- and perfusion-weighted imaging in healthy volunteers. Eur J Radiol. 2013;82(6):1015–22.

13. Zhao Q, Meng M, Kumar R, Wu Y, Huang J, Lian N, et al. The impact of COPD and smoking history on the severity of COVID-19: A systemic review and meta-analysis. J Med Virol. 2020.

14. Wu Z, McGoogan JM. Characteristics of and Important Lessons From the Coronavirus Disease 2019 (COVID-19) Outbreak in China: Summary of a Report of 72314 Cases From the Chinese Center for Disease Control and Prevention. JAMA. 2020.

15. Wei-jie Guan W-hL, Yi Zhao, Heng-rui Liang, Zi-sheng Chen, Yi-min Li, Xiao-qing Liu, Ru-chong Chen, Chun-li Tang, Tao Wang, Chun-quan Ou, Li Li, Ping-yan Chen, Ling Sang, Wei Wang, Jian-fu Li, Cai-chen Li, Li- min Ou, Bo Cheng, Shan Xiong, Zheng-yi Ni, Yu Hu, Jie Xiang, Lei Liu, Hong Shan, Chun-liang Lei, Yi-xiang Peng, Li Wei, Yong Liu, Ya-hua Hu, Peng Peng, Jian-ming Wang, Ji-yang Liu, Zhong Chen, Gang Li, Zhi-jian Zheng, Shao-qin Qiu, Jie Luo, Chang-jiang Ye, Shao-yong Zhu, Lin-ling Cheng, Feng Ye, Shi-yue Li, Jin-ping Zheng, Nuo-fu Zhang, Nan-shan Zhong, Jian-xing He. Comorbidity and its impact on 1,590 patients with COVID-19 in China: A Nationwide Analysis. medRxiv doi: https://doiorg/101101/2020022520027664.

16. Wichmann D, Sperhake JP, Lutgehetmann M, Steurer S, Edler C, Heinemann A, et al. Autopsy Findings and Venous Thromboembolism in Patients With COVID-19: A Prospective Cohort Study. Ann Intern Med. 2020.

17. Huang C, Wang Y, Li X, Ren L, Zhao J, Hu Y, et al. Clinical features of patients infected with 2019 novel coronavirus in Wuhan, China. Lancet. 2020;395(10223):497–506.

18. Ravi D, Wong C, Deligianni F, Berthelot M, Andreu-Perez J, Lo B, et al. Deep Learning for Health Informatics. IEEE J Biomed Health Inform. 2017;21(1):4–21.

19. Oelsner EC, Loehr LR, Henderson AG, Donohue KM, Enright PL, Kalhan R, et al. Classifying Chronic Lower Respiratory Disease Events in Epidemiologic Cohort Studies. Ann Am Thorac Soc. 2016;13(7):1057–66.

20. The Atherosclerosis Risk in Communities (ARIC) Study: design and objectives. The ARIC investigators. Am J Epidemiol. 1989;129(4):687–702.

21. Friedman GD, Cutter GR, Donahue RP, Hughes GH, Hulley SB, Jacobs DR, Jr., et al. CARDIA: study design, recruitment, and some characteristics of the examined subjects. J Clin Epidemiol. 1988;41(11):1105–16.

22. Tsao CW, Vasan RS. Cohort Profile: The Framingham Heart Study (FHS): overview of milestones in cardiovascular epidemiology. Int J Epidemiol. 2015;44(6):1800–13.

23. Daviglus ML, Talavera GA, Aviles-Santa ML, Allison M, Cai J, Criqui MH, et al. Prevalence of major cardiovascular risk factors and cardiovascular diseases among Hispanic/Latino individuals of diverse backgrounds in the United States. JAMA. 2012;308(17):1775–84.

24. Lavange LM, Kalsbeek WD, Sorlie PD, Aviles-Santa LM, Kaplan RC, Barnhart J, et al. Sample design and cohort selection in the Hispanic Community Health Study/Study of Latinos. Ann Epidemiol. 2010;20(8):642–9.

25. Sorlie PD, Aviles-Santa LM, Wassertheil-Smoller S, Kaplan RC, Daviglus ML, Giachello AL, et al. Design and implementation of the Hispanic Community Health Study/Study of Latinos. Ann Epidemiol. 2010;20(8):629–41.

26. Carpenter MA, Crow R, Steffes M, Rock W, Heilbraun J, Evans G, et al. Laboratory, reading center, and coordinating center data management methods in the Jackson Heart Study. Am J Med Sci. 2004;328(3):131–44.

27. Keku E, Rosamond W, Taylor HA, Jr., Garrison R, Wyatt SB, Richard M, et al. Cardiovascular disease event classification in the Jackson Heart Study: methods and procedures. Ethn Dis. 2005;15(4 Suppl 6):S6-62-70.

28. Taylor HA, Jr., Wilson JG, Jones DW, Sarpong DF, Srinivasan A, Garrison RJ, et al. Toward resolution of cardiovascular health disparities in African Americans: design and methods of the Jackson Heart Study. Ethn Dis. 2005;15(4 Suppl 6):S6-4-17.

29. Kanaya AM, Chang A, Schembri M, Puri-Taneja A, Srivastava S, Dave SS, et al. Recruitment and retention of US South Asians for an epidemiologic cohort: Experience from the MASALA study. J Clin Transl Sci. 2019;3(2-3):97–104.

30. Kanaya AM, Kandula N, Herrington D, Budoff MJ, Hulley S, Vittinghoff E, et al. Mediators of Atherosclerosis in South Asians Living in America (MASALA) study: objectives, methods, and cohort description. Clin Cardiol. 2013;36(12):713–20.

31. Bild DE, Bluemke DA, Burke GL, Detrano R, Diez Roux AV, Folsom AR, et al. Multi-Ethnic Study of Atherosclerosis: objectives and design. Am J Epidemiol. 2002;156(9):871–81.

32. Lee ET, Welty TK, Fabsitz R, Cowan LD, Le NA, Oopik AJ, et al. The Strong Heart Study. A study of cardiovascular disease in American Indians: design and methods. Am J Epidemiol. 1990;132(6):1141–55.

33. North KE, Howard BV, Welty TK, Best LG, Lee ET, Yeh JL, et al. Genetic and environmental contributions to cardiovascular disease risk in American Indians: the strong heart family study. Am J Epidemiol. 2003;157(4):303–14.

34. Regan EA, Hokanson JE, Murphy JR, Make B, Lynch DA, Beaty TH, et al. Genetic epidemiology of COPD (COPDGene) study design. COPD. 2010;7(1):32–43.

35. Couper D, LaVange LM, Han M, Barr RG, Bleecker E, Hoffman EA, et al. Design of the Subpopulations and Intermediate Outcomes in COPD Study (SPIROMICS). Thorax. 2014;69(5):491–4.

36. Teague WG, Phillips BR, Fahy JV, Wenzel SE, Fitzpatrick AM, Moore WC, et al. Baseline Features of the Severe Asthma Research Program (SARP III) Cohort: Differences with Age. J Allergy Clin Immunol Pract. 2018;6(2):545–54 e4.

37. Sacco RL, Boden-Albala B, Gan R, Chen X, Kargman DE, Shea S, et al. Stroke incidence among white, black, and Hispanic residents of an urban community: the Northern Manhattan Stroke Study. Am J Epidemiol. 1998;147(3):259–68.

38. Howard VJ, Cushman M, Pulley L, Gomez CR, Go RC, Prineas RJ, et al. The reasons for geographic and racial differences in stroke study: objectives and design. Neuroepidemiology. 2005;25(3):135–43.

39. Psaty BM, O’Donnell CJ, Gudnason V, Lunetta KL, Folsom AR, Rotter JI, et al. Cohorts for Heart and Aging Research in Genomic Epidemiology (CHARGE) Consortium: Design of prospective meta-analyses of genome-wide association studies from 5 cohorts. Circ Cardiovasc Genet. 2009;2(1):73–80.

40. Oelsner EC, Balte PP, Cassano PA, Couper D, Enright PL, Folsom AR, et al. Harmonization of Respiratory Data From 9 US Population-Based Cohorts: The NHLBI Pooled Cohorts Study. Am J Epidemiol. 2018;187(11):2265–78.

41. Cross Cohort Collaboration [Available from: https://chs-nhlbi.org/node/6539.

42. Levine DA, Gross AL, Briceno EM, Tilton N, Kabeto MU, Hingtgen SM, et al. Association Between Blood Pressure and Later-Life Cognition Among Black and White Individuals. JAMA Neurol. 2020;77(7):810–9.

43. Taliun D, Harris DN, Kessler MD, Carlson J, Szpiech ZA, Torres R, et al. Sequencing of 53,831 diverse genomes from the NHLBI TOPMed Program. Nature. 2021;590(7845):290–9.

44. Flanagin A, Frey T, Christiansen SL, Bauchner H. The Reporting of Race and Ethnicity in Medical and Science Journals: Comments Invited. JAMA. 2021.

45. Oelsner MESA COVID-19 Questionnaire 2020 [Available from: https://www.phenxtoolkit.org/toolkit_content/PDF/MESA_Questionnaire_Annotated.pdf.

46. (BRFSS) BRFSS. Questionnaire 2019 [Available from: https://www.cdc.gov/brfss/questionnaires/pdf-ques/2019-BRFSS-Questionnaire508.pdf.

47. (HRS) HaRS. COVID-19 Questionnaire 2020 [Available from: https://hrs.isr.umich.edu/sites/default/files/meta/2020/core/qnaire/online/05hr20COVID.pdf.

48. (MACS/WIHS-CSS) MACSWsIHSCCS. COVID-19 Questionnaire 2020 [Available from: https://www.phenxtoolkit.org/toolkit_content/PDF/MACSWIHS.pdf.

49. Andresen EM, Malmgren JA, Carter WB, Patrick DL. Screening for depression in well older adults: evaluation of a short form of the CES-D (Center for Epidemiologic Studies Depression Scale). Am J Prev Med. 1994;10(2):77–84.

50. Cohen S, Kamarck T, Mermelstein R. A global measure of perceived stress. J Health Soc Behav. 1983;24(4):385–96.

51. Levine DW, Kripke DF, Kaplan RM, Lewis MA, Naughton MJ, Bowen DJ, et al. Reliability and validity of the Women’s Health Initiative Insomnia Rating Scale. Psychol Assess. 2003;15(2):137–48.

52. Pilkonis PA, Choi SW, Reise SP, Stover AM, Riley WT, Cella D, et al. Item banks for measuring emotional distress from the Patient-Reported Outcomes Measurement Information System (PROMIS(R)): depression, anxiety, and anger. Assessment. 2011;18(3):263–83.

53. RAND. Social Support Survey Instrument [Available from: https://www.rand.org/health/surveys_tools/mos/social-support/surveyinstrument.html.

54. Russell DW. UCLA Loneliness Scale (Version 3): reliability, validity, and factor structure. J Pers Assess. 1996;66(1):20–40.

55. Smith BW, Dalen J, Wiggins K, Tooley E, Christopher P, Bernard J. The brief resilience scale: assessing the ability to bounce back. Int J Behav Med. 2008;15(3):194–200.

56. Harris PA, Taylor R, Minor BL, Elliott V, Fernandez M, O’Neal L, et al. The REDCap consortium: Building an international community of software platform partners. J Biomed Inform. 2019;95:103208.

57. Harris PA, Taylor R, Thielke R, Payne J, Gonzalez N, Conde JG. Research electronic data capture (REDCap)--a metadata-driven methodology and workflow process for providing translational research informatics support. J Biomed Inform. 2009;42(2):377–81.

58. CMS Guidelines for COVID-19 Coding 2020 [Available from: https://www.cdc.gov/nchs/data/icd/COVID-19-guidelines-final.pdf.

59. Zakai NA, McClure LA, Judd SE, Safford MM, Folsom AR, Lutsey PL, et al. Racial and regional differences in venous thromboembolism in the United States in 3 cohorts. Circulation. 2014;129(14):1502–9.

60. CDC. Interim Guidelines for COVID-19 Antibody Testing [Available from: https://www.cdc.gov/coronavirus/2019-ncov/lab/resources/antibody-tests-guidelines.html#anchor_1590264293982.

61. Laing ED, Sterling SL, Richard SA, Epsi NJ, Coggins S, Samuels EC, et al. Antigen-based multiplex strategies to discriminate SARS-CoV-2 natural and vaccine induced immunity from seasonal human coronavirus humoral responses. medRxiv. 2021.

62. Balte PP, Chaves PHM, Couper DJ, Enright P, Jacobs DR, Jr., Kalhan R, et al. Association of Nonobstructive Chronic Bronchitis With Respiratory Health Outcomes in Adults. JAMA Intern Med. 2020.

63. Bhatt SP, Balte PP, Schwartz JE, Cassano PA, Couper D, Jacobs DR, Jr., et al. Discriminative Accuracy of FEV1:FVC Thresholds for COPD-Related Hospitalization and Mortality. JAMA. 2019;321(24):2438–47.

64. Bhatt SP, Schwartz JE, Oelsner EC. FEV1:FVC Thresholds for Defining Chronic Obstructive Pulmonary Disease-Reply. JAMA. 2019;322(16):1611–2.

65. Cornelius T BP, Bhatt SP, Cassano PA, Currow D, Jacobs DR, Johnson M, Kalhan R, Kronmal R, Loehr LR, O’Connor GT, Schwartz JE, Smith BM, White WB, Yende S, Oelsner EC. A dyadic growth modeling approach to weight gain and lung function loss: The NHLBI Pooled Cohorts Study. Am J Epidemiol. 2020 (Accepted).

66. Oelsner EC, Balte PP, Bhatt SP, Cassano PA, Couper D, Folsom AR, et al. Lung function decline in former smokers and low-intensity current smokers: a secondary data analysis of the NHLBI Pooled Cohorts Study. Lancet Respir Med. 2020;8(1):34–44.

67. Zhang Y, Vittinghoff E, Pletcher MJ, Allen NB, Zeki Al Hazzouri A, Yaffe K, et al. Associations of Blood Pressure and Cholesterol Levels During Young Adulthood With Later Cardiovascular Events. J Am Coll Cardiol. 2019;74(3):330–41.

68. Levine DA, Gross AL, Briceno EM, Tilton N, Kabeto MU, Hingtgen SM, et al. Association Between Blood Pressure and Later-Life Cognition Among Black and White Individuals. JAMA Neurol. 2020.

69. Griffith L, van den Heuvel E, Fortier I, Hofer S, Raina P, Sohel N, Payette H, Wolfson C, Belleville S. Harmonization of Cognitive Measures in Individual Participant Data and Aggregate Data Meta-Analysis. Methods Research Report. (Prepared by the McMaster University Evidence-based Practice Center under Contract No. 290-2007-10060-I.) AHRQ Publication No.13-EHC040-EF. Rockville, MD: Agency for Healthcare Research and Quality; March 2013. www.effectivehealthcare.ahrq.gov/reports/final.cfm.

70. Griffith LE, van den Heuvel E, Fortier I, Sohel N, Hofer SM, Payette H, et al. Statistical approaches to harmonize data on cognitive measures in systematic reviews are rarely reported. J Clin Epidemiol. 2015;68(2):154–62.

71. Gross AL, Jones RN, Fong TG, Tommet D, Inouye SK. Calibration and validation of an innovative approach for estimating general cognitive performance. Neuroepidemiology. 2014;42(3):144–53.

72. Gross AL, Sherva R, Mukherjee S, Newhouse S, Kauwe JS, Munsie LM, et al. Calibrating longitudinal cognition in Alzheimer’s disease across diverse test batteries and datasets. Neuroepidemiology. 2014;43(3- 4):194–205.

73. Gross AL, Mungas DM, Crane PK, Gibbons LE, MacKay-Brandt A, Manly JJ, et al. Effects of education and race on cognitive decline: An integrative study of generalizability versus study-specific results. Psychol Aging. 2015;30(4):863–80.

74. Langa KM, Ryan LH, McCammon RJ, Jones RN, Manly JJ, Levine DA, et al. The Health and Retirement Study Harmonized Cognitive Assessment Protocol Project: Study Design and Methods. Neuroepidemiology. 2020;54(1):64–74.

75. Oelsner EC, Ortega VE, Smith BM, Nguyen JN, Manichaikul AW, Hoffman EA, et al. A Genetic Risk Score Associated with Chronic Obstructive Pulmonary Disease Susceptibility and Lung Structure on Computed Tomography. Am J Respir Crit Care Med. 2019;200(6):721–31.

76. Smith BM, Kirby M, Hoffman EA, Kronmal RA, Aaron SD, Allen NB, et al. Association of Dysanapsis With Chronic Obstructive Pulmonary Disease Among Older Adults. JAMA. 2020;323(22):2268–80.

77. Smith BM, Hoffman EA, Rabinowitz D, Bleecker E, Christenson S, Couper D, et al. Comparison of spatially matched airways reveals thinner airway walls in COPD. The Multi-Ethnic Study of Atherosclerosis (MESA) COPD Study and the Subpopulations and Intermediate Outcomes in COPD Study (SPIROMICS). Thorax. 2014;69(11):987–96.

78. Hame Y, Angelini E, Hoffman E, Barr G, Laine A. Adaptive quantification and longitudinal analysis of pulmonary emphysema with a hidden Markov measure field model. IEEE Transactions on Medical Imaging. 2014;33(7):1527–40.

79. Wang M, Aaron CP, Madrigano J, Hoffman EA, Angelini E, Yang J, et al. Association Between Long-term Exposure to Ambient Air Pollution and Change in Quantitatively Assessed Emphysema and Lung Function. JAMA. 2019;322(6):546–56.

80. Yang J, Angelini E, Smith BM, Balte P, Hoffman E, Barr RG, et al. Unsupervised Discovery of Spatially-Informed Lung Texture Patterns for Pulmonary Emphysema: The MESA COPD Study. International Conference on Medical Image Computing and Computer-Assisted Intervention (MICCAI)2017. p. 116–24.

81. Yang J, Angelini ED, Balte PP, Hoffman EA, Wu CO, Venkatesh BA, et al. Emphysema quantification on cardiac CT scans using hidden Markov measure field model: The MESA Lung Study. International Conference on Medical Image Computing and Computer-Assisted Intervention (MICCAI); Athenes, Greece: Springer; 2016. p. 624–31.

82. Yang J, Vetterli T, Balte PP, Barr RG, Laine AF, Angelini ED. Unsupervised Domain Adaption With Adversarial Learning (UDAA) for Emphysema Subtyping on Cardiac CT Scans: The Mesa Study. IEEE International Symposium on Biomedical Imaging (ISBI)2019. p. 289–93.

83. Pham T TT, Phung D and Venkatesh S.. Deepcare: A deep dynamic memory model for predictive medicine. Paper presented at: Pacific-Asia Conference on Knowledge Discovery and Data Mining; 2016.

84. Miotto R, Li L, Kidd BA, Dudley JT. Deep Patient: An Unsupervised Representation to Predict the Future of Patients from the Electronic Health Records. Sci Rep. 2016;6:26094.

85. He K, ZHang X, Ren S, Sun J. (2015-12-10). “Deep Residual Learning for Image Recognition”. arXiv:1512.03385.

86. Brody JA, Morrison AC, Bis JC, O’Connell JR, Brown MR, Huffman JE, et al. Analysis commons, a team approach to discovery in a big-data environment for genetic epidemiology. Nat Genet. 2017;49(11):1560–3.

87. Health NIo. NIH Tribal Consultation Report: NIH Draft Policy for Data Management and Sharing 2020 [Available from: https://osp.od.nih.gov/wp-content/uploads/Tribal_Report_Final_508.pdf.

88. Roger VL, Boerwinkle E, Crapo JD, Douglas PS, Epstein JA, Granger CB, et al. Strategic transformation of population studies: recommendations of the working group on epidemiology and population sciences from the National Heart, Lung, and Blood Advisory Council and Board of External Experts. Am J Epidemiol. 2015;181(6):363–8.

